# Impact of climate change on diarrhoea risk in low- and middle-income countries

**DOI:** 10.1101/2024.12.24.24319610

**Authors:** Syeda H. Fatima, Melinda A. Judge, Peter N. Le Souëf, Corey J. A. Bradshaw

## Abstract

Diarrhoea remains a leading cause of mortality among children under five years of age, with over 99% of deaths occurring in low- and middle-income countries. Poor water quality, inadequate sanitation, poverty, undernutrition, and limited healthcare access contribute to this lingering problem, along with emerging environmental stressors driven by climate change. We analysed long-term spatial relationships between environmental, socioeconomic, and maternal/child factors using *Demographic and Health Surveys* and *WorldClim* data across eight South and Southeast Asian countries (*n* = 66,545 clusters; 3,143,811 child-level observations).

We employed boosted regression trees to assess variable importance across five thematic phases: socio-economic, maternal, child, climate, and combined. We selected variables based on biological plausibility, collinearity checks, and completeness. We addressed uncertainty through multiple imputation and stochastic resampling, and we evaluated model performance using cross-validation.

The main predictors of diarrhoea incidence included annual temperature variability, precipitation in the wettest month, maternal education, and household size. Higher annual temperature range (30–40 °C) was associated with a ∼ 39% increase in diarrhoea probability, while lower precipitation in the wettest month (< 600 mm) increased risk by ∼ 29%, highlighting the role of drier conditions. We found that maternal education < 8 years increased diarrhoea probability by ∼ 18%, and household sizes exceeding six members increased it by ∼ 9%.

Our findings emphasise the need for climate-resilient public-health policies that integrate social and environmental determinants of diarrhoea. Targeted interventions — including improved maternal education, water and sanitation infrastructure, and resource management in densely populated households — are necessary to mitigate diarrhoea risk in vulnerable regions under changing climate conditions.

## Introduction

Diarrhoea remains a leading cause of death among children under five years of age worldwide, causing an estimated 443,833 deaths annually (World Health Organization, 2024). Although this marks a decrease from the 1,197,044 diarrhoea-related deaths in children under five in 2000, it remains a major global health challenge (Arifin et al., 2022; Black et al., 2024; Das et al., 2024). In addition to contributing to child mortality, repeated bouts of acute diarrhoea, as well as chronic and persistent diarrhoea, have long-term health consequences, including undernutrition, weakened immune systems, impaired cognitive development, and a higher risk of cardiometabolic diseases in adulthood (Das et al., 2024; DeBoer et al., 2013). Over 99% of diarrhoea mortality in under-five children occurs in low- and middle-income countries (Black et al., 2024). Sub-Saharan Africa and Southeast Asia bear most of the global diarrhoeal burden, accounting for > 80% of under-five deaths from the disease (Troeger et al., 2018; World Health Organization, 2022).

Many interconnected factors drive the high incidence of diarrhoea in low- and middle-income countries, including poor-quality drinking water, inadequate sanitation, unsanitary hygiene conditions, poverty, undernutrition, limited access to healthcare, high population density, lack of education, and other environmental drivers. The lack of clean water and adequate sanitation often leads to the consumption of contaminated water and food, while poor hygiene practices further spread diarrhoeal diseases (Gebrezgiabher et al., 2019; Getachew et al., 2018; McClelland et al., 2022; Roushdy et al., 2012; Tafere et al., 2020; Workie et al., 2019). Around 88% of diarrhoeal deaths are linked to unsafe drinking water and related causes (Centers for Disease Control and Prevention, 2024). One study found that improved access to drinking water reduced the risk of diarrhoea by 52%, while improved sanitation facilities lowered the risk by 24% (Wolf et al., 2022). Poverty increases the risk of diarrhoea by limiting access to nutritious food, clean water, and healthcare, while promoting environments where diarrhoeal pathogens thrive (Woldu et al., 2016).

Childhood undernutrition and infectious disease are intricately linked in a cyclical relationship, with each being both a risk factor and an outcome of the other. Undernutrition weakens the immune system, making children more vulnerable to infections such as diarrhoea, which in turn leads to further nutrient loss and impaired recovery (Morales et al., 2023; Siddiqui et al., 2021; Wasihun et al., 2018). For example, diarrhoea can exacerbate undernutrition by reducing food intake, nutrient absorption, and growth, while undernourished children are slower to recover, increasing the risk of severe dehydration and death. Once started, this cycle can become self-exacerbating. Furthermore, healthcare facilities in many low- and middle-income countries are often under-resourced and difficult to access, especially in rural areas, preventing children from receiving timely or adequate treatment: oral rehydration therapy essential for reducing dehydration-related fatalities (Walker et al., 2012).

Rapid urbanisation, overcrowding, and high population densities in low- and middle-income countries also facilitate the transmission of diarrhoeal pathogens, including bacteria (*Escherichia coli*, *Shigella* spp., *Salmonella*, *Campylobacter* spp., *Vibrio cholerae*), viruses (*Rotavirus*, *Norovirus*, *Adenovirus, Astrovirus*, *Sapovirus*), and parasites (*Giardia lamblia*, *Cryptosporidium* spp., *Entamoeba histolytica*) (Bishwajit et al., 2014; Black et al., 2024; Tosam et al., 2019). A lack of education compounds the issue, limiting awareness of good hygiene practices, the importance of breastfeeding, and recognising the symptoms of diarrhoea-related dehydration — all necessary for preventing and managing diarrhoea (Desmennu et al., 2017). High temperatures and precipitation further increase the spread of diarrhoea-causing pathogens (Geremew et al., 2024; Liang et al., 2021; Wibawa et al., 2023; Xu et al., 2013). Indeed, higher temperatures are associated with an increased incidence of diarrhoea, especially those caused by bacteria, while heavy rainfall after dry periods has been linked to higher diarrhoea prevalence (Carlton et al., 2014). Diarrhoea might also increase during the dry season due to water scarcity, leading to a reliance of unsafe water sources and reduced hygiene practices (Wang et al., 2022).

Climate change is expected to worsen these challenges by increasing the frequency of extreme weather events (especially catastrophic flooding; (Hirabayashi et al., 2013; Trancoso et al., 2024)), disrupting food security (Myers et al., 2017), undermining social safety nets (Hallegatte & Rozenberg, 2017; Thomas et al., 2019), and reducing access to healthcare facilities (Romanello et al., 2024), thereby escalating the diarrhoea burden in vulnerable populations (Alexander et al., 2013; Levy et al., 2016). For example, the 2022 floods in Pakistan and cyclone Amphan in Bangladesh and India caused severe damage, including contamination of water resources, destruction of sanitation facilities, and widespread displacement, thereby increasing the incidence of diarrhoea and other infectious diseases (Manzoor & Adesola, 2022; Rafa et al., 2021).

Despite the public health impact, few studies have comprehensively examined the combined effects of socio-economic, behavioural, and environmental determinants of diarrhoeal incidence and risk, particularly in Southeast Asia where diarrhoea in young children is pervasive. Additionally, diarrhoea-prevention programs are not always implemented uniformly, reducing their effectiveness in areas most in need. Studies that assess the aetiology of diarrhoea at fine spatial resolutions can provide detailed evidence to guide local-scale policies and interventions, maximising their impact where they are most needed (Evans et al., 2021; Evans et al., 2024; Pourtois et al., 2023).

In this study we aimed to determine the most important predictors of diarrhoea in children under five in South and Southeast Asia by exploring the long-term spatial associations between diarrhoeal incidence and various behavioural, socio-demographic, and environmental factors. Our focus is on providing quantifiable evidence to identify the most efficient interventions, enabling strategies to target location-specific risk factors effectively. We hypothesise that the incidence of diarrhoea in children under five varies across locations and time, influenced by a combination of factors including the child’s and mother’s health (e.g., nutritional status, anaemia, body-mass index, vaccinations), the family’s socio-economic conditions (income, education, access to clean water, sanitation, and healthcare), hygiene practices (child stool disposal), and environmental conditions (annual temperature, temperature range, precipitation). We expect this variation to highlight distinct regional risk profiles, necessitating tailored interventions to reduce diarrhoea incidence.

## Methods

We used data on diarrhoea and other individual and household characteristics from the Demographic and Health Surveys (dhsprogram.com) women’s birth history (BR) data for children under the age of five years. The Demographic and Health Surveys program has organised nationally representative household surveys in low- and middle-income countries since 1984, covering a wide range of topics such as family planning, maternal and child health, malaria, nutrition, and environmental health. The burden of childhood diarrhoea remains high in South Asian and Southeast Asian. Specifically, we included 17 available surveys with 3,143,811 observations from eight countries in South Asia and Southeast Asia (Fig. 1). To ensure consistency and avoid discrepancies from earlier versions, we used surveys done from 2010 onwards, using only Demographic and Health Survey phases 6, 7, and 8, which also provided global positioning data. Supplementary data table S1 provides the details of countries, year of surveys, and global positioning data that we included in the analysis. We obtained ethics approval from the Flinders University Human Research Ethics Committee (HEG8026-2).

**Figure 1.**
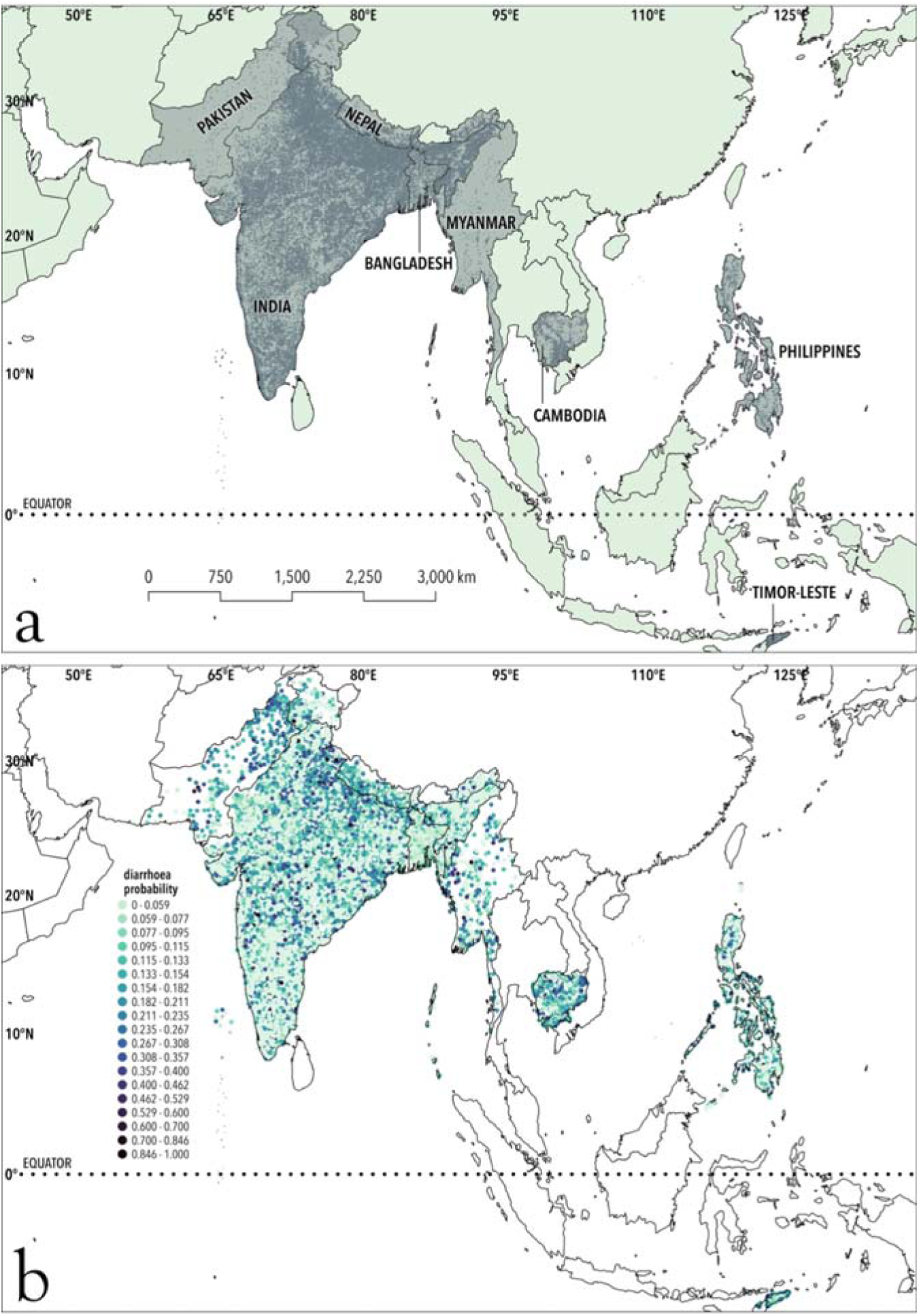
(a) Map highlighting the eight countries in South and Southeast Asia that we included in the analysis. (b) Probability distribution of diarrhoea cases at the cluster level across the eight countries in the study region.

We estimated each child’s age in months by calculating the difference between the interview date (V008) and the birth date (B3). The final dataset included only living children under the age of five (≤ 60 months; *n* = 593,315).

Our hypothesis is that diarrhoea incidence among children under five is influenced by a complex interplay of child characteristics, maternal characteristics, socio-economic conditions, and environmental factors. To test these hypotheses, we selected variables based on three criteria: (*i*) their plausible association with diarrhoea incidence as supported by published evidence; (*ii*) an assessment of collinearity with other variables; and (*iii*) the extent of missing data for each variable (< 25%).

We did a literature review to identify causal associations. Evidence suggests that child health indicators, including nutritional status, anaemia, birth weight, and drugs for intestinal parasites, influence susceptibility to diarrhoea (Morales et al., 2023; Siddiqui et al., 2021; Wasihun et al., 2018). Maternal health indicators including mother’s age at birth, body mass index, and delivery by caesarean section, can influence long-term infant outcomes and are potentially associated with the incidence of diarrhoea (Althaus et al., 2022; Finlay et al., 2011; Neu & Rushing, 2011). Socio-economic factors, including household wealth, maternal education, access to health care, clean water, and sanitation facilities, are determinants of health outcomes, both impacting risk and severity of diarrhoea (Desmennu et al., 2017; Gebrezgiabher et al., 2019; Woldu et al., 2016; Workie et al., 2019). Environmental conditions, particularly mean annual temperature, annual temperature range, extremes, mean precipitation, and precipitation seasonality, are also associated with diarrhoea incidence, especially in the context of climate change (Geremew et al., 2024; Liang et al., 2021; Wibawa et al., 2023; Xu et al., 2013).

These diverse risk factors underscore the importance of location-specific interventions to reduce diarrhoea and improve child health outcomes. Supplementary data table S2 provides details on the list of variables we included and their recoded values. Supplementary data table S3 provides information on missing data in the final selected variables.

To account for missing data, we applied multiple imputation. Initially, we identified variables with missing values and categorised them into categorical and continuous types. We then converted categorical variables to factors and continuous variables to numeric. Using the `mice` package (Van Buuren & Groothuis-Oudshoorn, 2011) in R (R Core Team 2024), we applied polytomous regression for categorical variables and predictive mean matching for continuous variables. We specified the imputation methods for each variable type, and then merged the imputed values into the original dataset.

We handled special cases such as *anemia_level*, *received_rotavirus*, and *region* by filtering specific values and using relevant predictors for imputation. We excluded *received_rotavirus* from the analysis because rotavirus vaccines were introduced after 2018 in most South and Southeast Asian countries, so a large proportion of dataset was rendered ineligible. We created histograms to visualise variable distributions before and after imputation (Supplementary data figure S1).

### Data aggregation at cluster level

We summarised the household level data at the cluster level (*n* = 66,545) by processing different variable types — continuous, binary, and categorical — using appropriate statistical methods. For continuous variables (*x*), we calculated both the mean μ*_x_* and variance σ^2^*_x_* at the cluster level. For binary variables, we assumed a Bernoulli distribution (Forbes et al., 2011) and estimated the probability *p* and variance σ^2^. For a binary variable, we calculated probability *p* as:

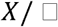

where *X* = the number of successes, and LJ = the number of observations. We then computed the variance

σ^2^ of the binary variables as:

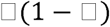

For ordinal categorical variables, we used the median at the cluster level to summarise the data. For climate data we used the WorldClim database (worldclim.org) at a spatial resolution of 30 arc-seconds. We used the geographic coordinates (longitude and latitude) of survey clusters to generate additional spatial points around each cluster. For each cluster, we supplemented a central point representing the true location by eight additional points located approximately 1.5 km away in cardinal and diagonal directions (Moore’s neighbourhood). For each cluster, we calculated the mean, variance, and standard deviation for all bioclimatic variables based on the central and its neighbouring points. We summarised these statistics across all 19 variables to represent the environmental variability within each cluster. From the 19 bioclimatic variables, we selected five: annual mean temperature (BIO1), temperature annual range (BIO7), annual precipitation (BIO12), precipitation seasonality (BIO15), and precipitation of the driest quarter (BIO17) based on their biological plausibility for assessing long-term trends, and low inter-correlation and variance inflation.

### Analysis

We applied boosted regression trees to explore the relationship between diarrhoea and various socio-economic, demographic, and climate-related factors according to our hypotheses. Boosted regression trees (machine learning) are well-suited to our data and objectives because the approach does not require a predefined model to build the regression trees as do traditional statistical methods. Instead, the approach applies an algorithm that learns the relationship between predictor variables (e.g., socio-economic factors) and the response variable (probability of diarrhoea), emphasising predictive performance rather than strict adherence to specified models.

The analysis followed several distinct thematic phases, with boosted regression trees applied to a set of selected variables in each phase. We began by focusing on socio-economic factors, including household size, wealth index, education, and access to healthcare. In subsequent phases, we analysed mother traits (e.g., maternal body mass index, maternal age, delivery by caesarean section), then child traits (e.g., gender, stunting, underweight, wasting), and finally climate variables (temperature annual range, temperature of the warmest month, precipitation in the wettest month, precipitation seasonality). For each phase, we created stochastic realisations of the data (resampling using Normal random deviates for continuous variables, binomial random deviates for binary variables, and multinomial random deviates for categorical variables) to account for variability and uncertainty.

We implemented boosted regression trees models using the *gbm.step* function from the `dismo` R package, setting a Gaussian distribution for the response variable (probability of diarrhoea). To optimise the models, we tuned several parameters, including the learning rate, tree complexity, and bag fraction. We applied cross-validation to assess model performance and to reduce overfitting. In each phase, we employed resampling techniques to ensure an equal number of samples from all countries, avoiding over- or under-representation and accounting for spatial autocorrelation.

For the final phase, we included the most influential variables from each thematic phase. We set the bag fraction to 0.5, the learning rate to 0.0001, the tolerance to 0.000001, and tree complexity to 2. To evaluate the goodness-of-fit of the final models, we used the cross-validation correlation coefficient (β_CV_), including its standard error calculated across all tree iterations.

We also implemented a kappa (κ) limitation to the resampled selections (Bradshaw & Brook, 2016). In this process, we retained only the resampled mean ranks within κ standard deviations (σ) of the overall average mean, with κ set to 2. After recalculating the mean and standard deviation of the ranks, we repeated this process several times. This iterative κσ ‘clipping’ approach, commonly used in image processing to remove artefacts, reduced the influence of outliers on the estimation of the mean rank across all 1000 iterations.

## Results

The total number of children < 5 years old at the household level was *n* = 593,315, and we analysed data at the cluster level (*n* = 66,545). Supplementary Table A1-3 gives descriptive statistics for prevalence of diarrhoea in children < 5 years old at the household level. Prevalence is generally higher in children between 6 and 23 months old. Among countries, diarrhoea prevalence in the 2 weeks preceding the survey was highly variable, ranging from 17.6% of under-fives in Pakistan to 4.85% in Bangladesh.

In thematic phase 1 (socio-economic variables), the most influential variables contributing to the variance in diarrhoea at a regional scale were women’s education and number of household members (24–26% relative influence) (Fig. 2a). Of these, women’s education had the highest relative contribution. In thematic phase 2 (maternal traits), postnatal care emerged as the most influential variable (30% relative influence), followed by maternal body-mass index and maternal age (Fig. 2b). In thematic phase 3 (child traits), drugs for intestinal parasites had the strongest influence (15%) (Fig. 2c). In thematic phase 4 (climate variables), annual temperature range and precipitation during the wettest month emerged as the most influential contributors (27–35%) (Fig. 2d). In the combined analysis (phase 5), the variables with the highest relative influence were annual temperature range, precipitation during the wettest month, women’s education, and number of household members (7–20%) (Fig. 2).

**Figure 2:**
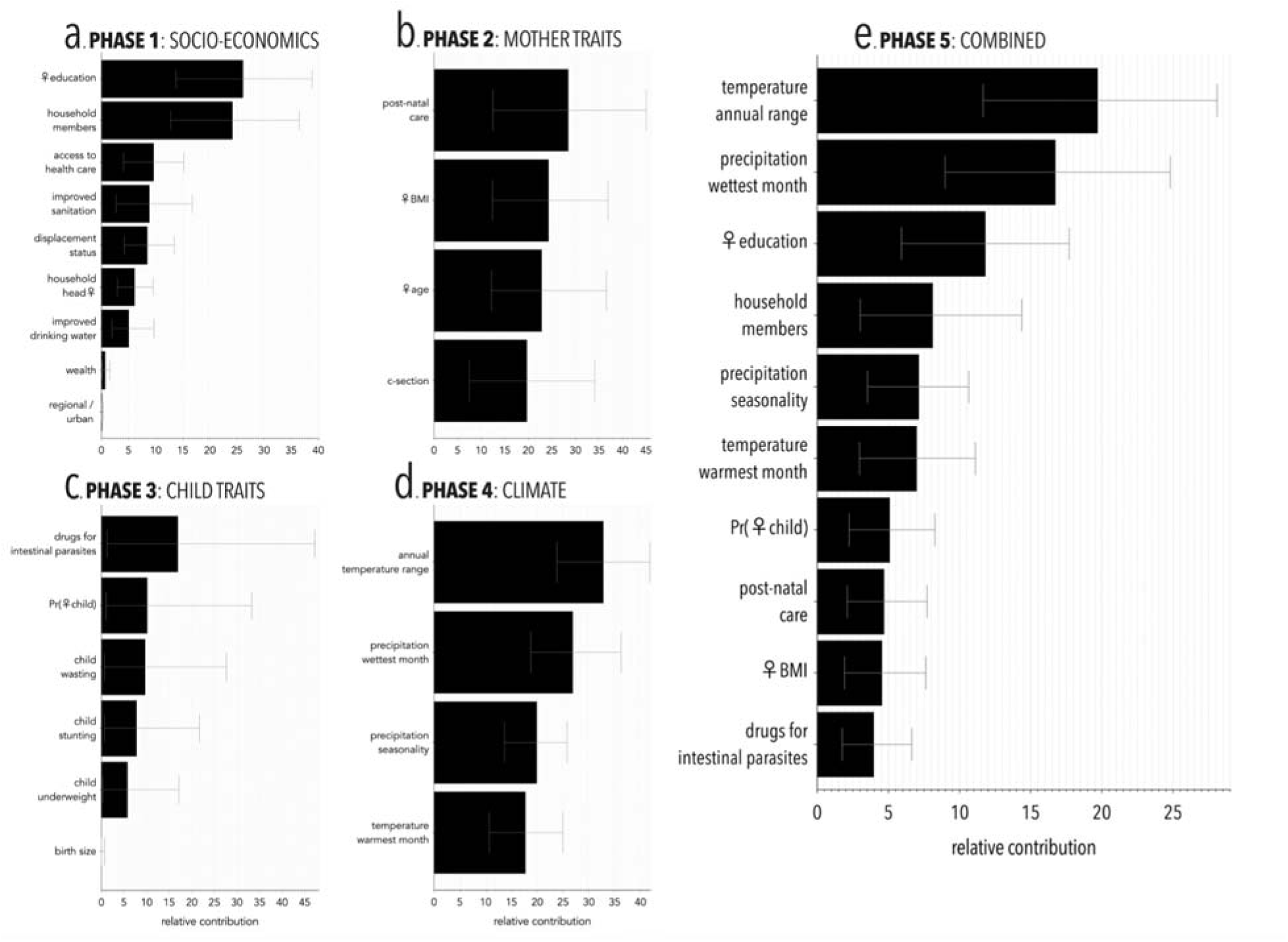
Relative influence of predictor variables on diarrhoea distribution across different model phases. (a) Socioeconomic factors, (b) maternal traits, (c) child traits, (d) climate variables, and (e) combined influence of the most influential variables from all phases.

The cross-validated deviance explained the final model’s goodness of fit in different phases, ranged between 6.70 and 14.37% for phase 1 (socio-economics), 1.42 and 8.23% for phase 2 (maternal traits), 1.10 and 6.90% for phase 3 (child traits), and 14.42 and 22.44% for phase 4 (climate). The combined phase 5 had a cross-validated deviance explained = 12.95–20.89%.

Figure 3 shows the response curves of the most influential variables contributing to the combined- phase boosted regression trees. An increase in the temperature annual range from approximately 30 °C to 40 °C is associated with a ∼ 39% increase in the probability of diarrhoea (Fig. 3a). Similarly, lower precipitation in the wettest month < 600 mm is associated with a ∼ 29% increase in the probability of diarrhoea (Fig. 3b). When a mother’s education falls below ∼ 8 years, there is an associated increase in the probability of diarrhoea by 18% as the number of years of education approaches zero (Fig. 3c). An increased number of household members beyond ∼ 6 is associated with a ∼ 9% increase in the probability of diarrhoea in children under five within those households (Fig. 3d).

**Figure 3:**
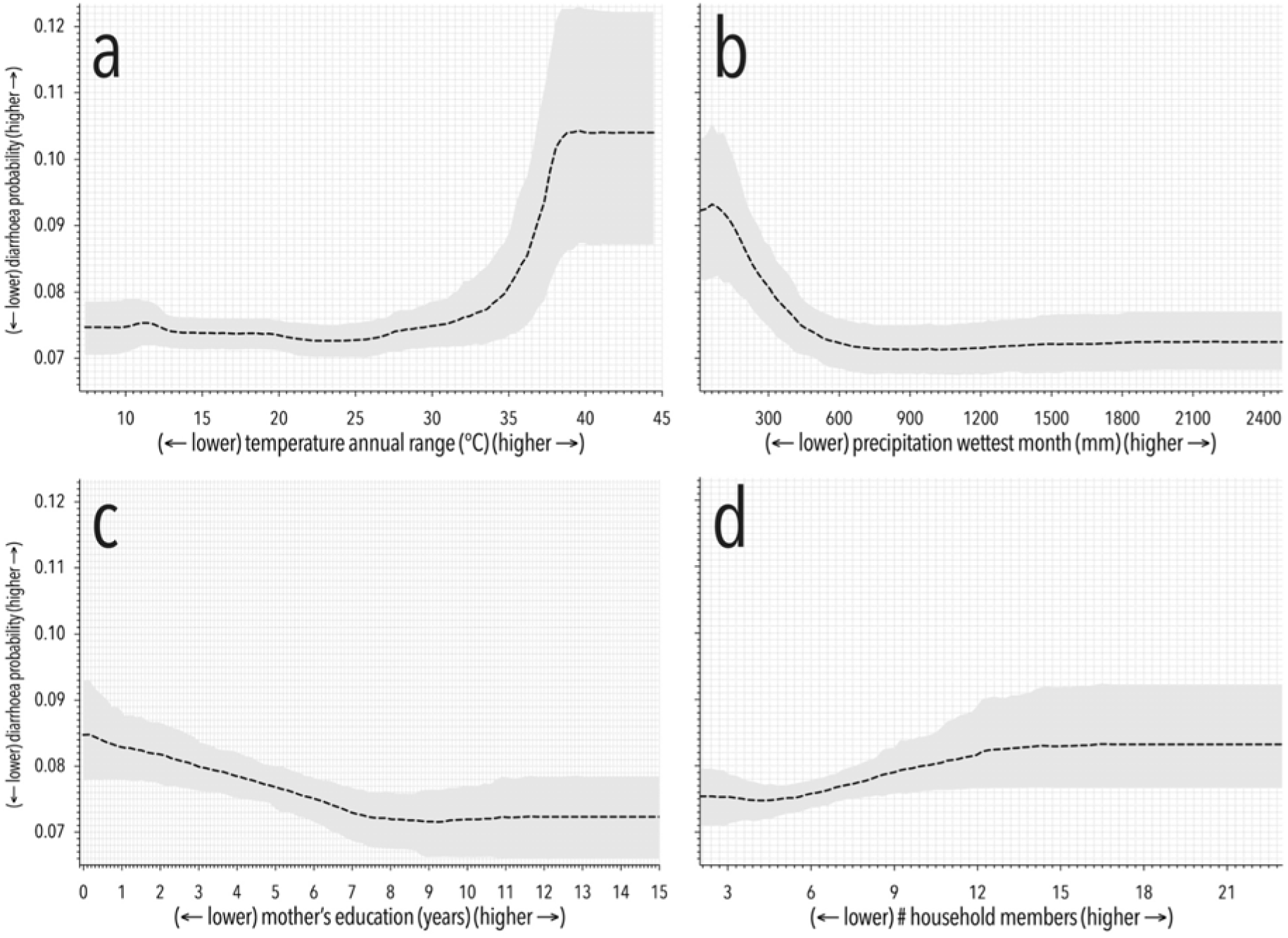
Predicted relationships between (a) temperature annual range, (b) precipitation in the wettest month, (c) mother’s education, and (d) number of household members on the probability of diarrhoea incidence.

Figure 4 shows the regional-scale spatial patterns of the main contributing variables in South and Southeast Asia. Annual temperature range indicates higher temporal variability in the inland subtropical regions of northern India and Pakistan, while the coastal tropical regions in southern India, Myanmar, Cambodia, and the Philippines exhibit a narrower temperature range (Fig. 4a). The tropical regions also experience higher precipitation during the wettest month (Fig. 4b). Women’s education is relatively higher in southern India, Bangladesh, and the Philippines (Fig. 4c). Darker shades indicate areas with larger average household sizes, predominantly clustered in northern India, western Pakistan, and parts of the Philippines (Fig. 4d).

**Figure 4.**
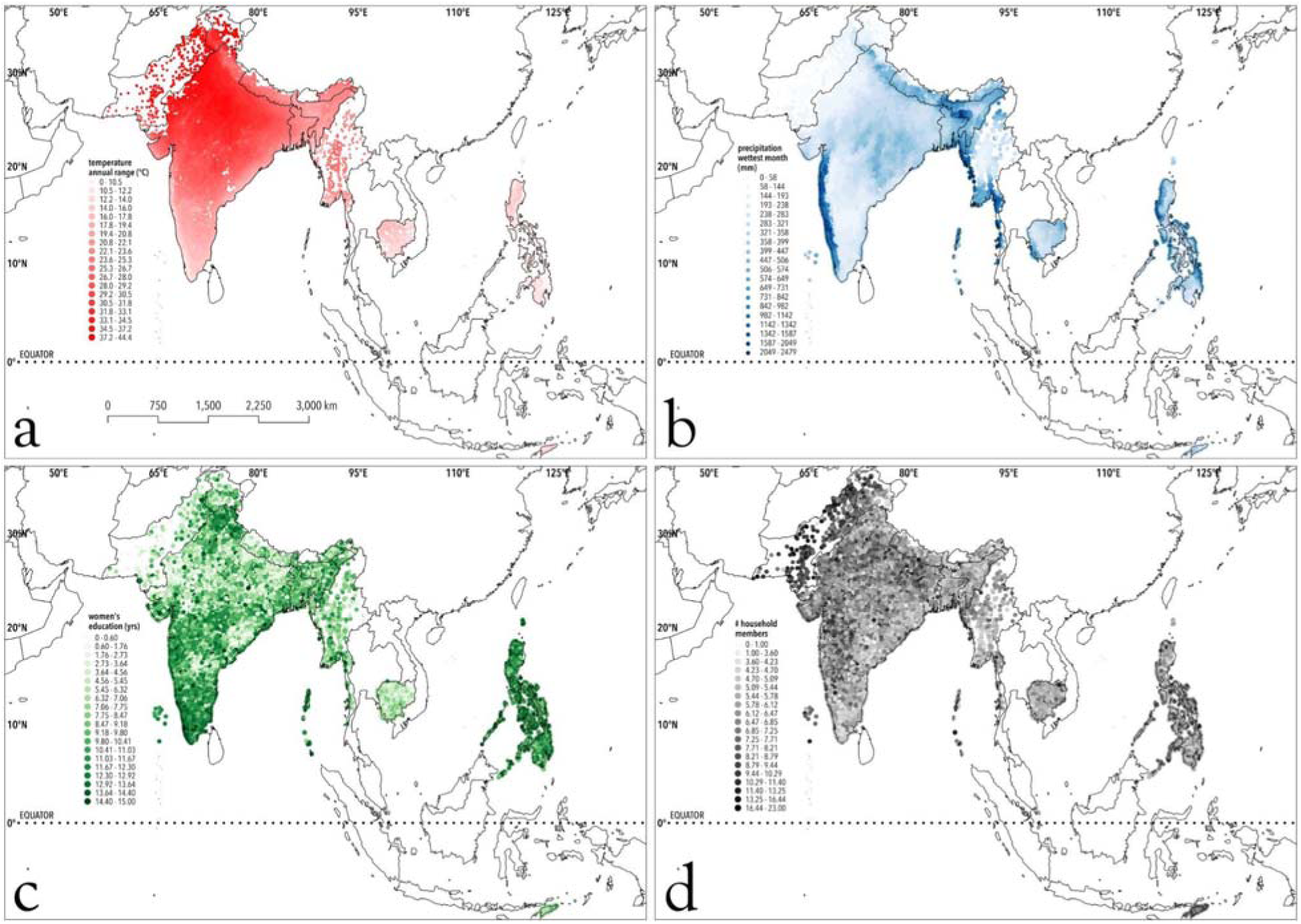
Spatial patterns of the most influential variables on the probability of diarrhoea: (a) temperature annual range, (b) precipitation of the wettest month, (c) mother’s education, (d) household members

## Discussion

The most influential predictors of diarrhoea incidence in South and Southeast Asia include annual temperature variability, precipitation in the wettest month, maternal education, and household size. The boosted regression tree response curves reveal that higher annual temperature variability correlates with a heightened probability of diarrhoea, whereas higher precipitation during peak rainfall months is associated with a reduced probability, underscoring the impact of drier conditions on disease spread in the region. As expected, we found evidence that higher maternal education correlates with decreased incidence of diarrhoea, while larger household sizes are linked to increased risk, possibly due to diluted resources and increased risk of spread of infections in the latter. Other variables had considerably less influence to explain regional variance in diarrhoea incidence.

The effects of daily extreme temperatures, diurnal variation, seasonality, and extreme weather events on the dynamics of diarrhoea are well-documented (Altizer et al., 2006; Bandyopadhyay et al., 2012; Chao et al., 2019; Lal et al., 2012; Liang et al., 2021; Pascual et al., 2008). Recently, research has delved further into the role of short-term temperature variability in influencing diarrhoeal infections, as well as overall morbidity and mortality (Gasparrini et al., 2015; Musengimana et al., 2016). However, emerging research now suggests a health risk posed by higher annual temperature variability on mortality and morbidity (Guo et al., 2021; Healy et al., 2023; Shi et al., 2015; Shi et al., 2016; Zanobetti et al., 2012). Despite these findings, the long-term impact of annual temperature variability on infectious disease incidence — particularly diarrhoea — remains an underexplored area. Our study addresses this gap by demonstrating that annual temperature variability is a predictor of diarrhoea outcomes at the regional scale, underscoring its potential to escalate disease burden as climate change-driven variability increases (Intergovernmental Panel on Climate Change., 2022).

Climate projections indicate that seasonal temperature ranges are likely to become more variable due to climate-driven extreme events, such as more frequent heatwaves and sporadic cold snaps (Intergovernmental Panel on Climate Change., 2022; van der Wiel & Bintanja, 2021). South and Southeast Asian countries, already vulnerable to higher temperature variability (Olonscheck et al., 2021), could face heightened risk of diarrhoea as this increased intra-annual variability exacerbates water scarcity, promotes pathogen growth in water and food, limits hygiene due to reduced access to clean water, weakens immune resilience, and strains sanitation infrastructure. Temperature fluctuations affect the ecology of diarrheal pathogens, particularly water-borne and food-borne organisms such as *Vibrio cholerae* and *E. coli* (Dietrich et al., 2023; Levy et al., 2016). Warmer conditions promote bacterial proliferation and persistence in water sources, elevating contamination risks. In regions with marked seasonal temperature shifts, diarrhoea prevalence often spikes in warmer months due to increased bacterial activity (Lal et al., 2012). This seasonal disease pattern can strain healthcare resources and contribute to outbreaks, particularly in communities lacking sufficient water-sanitation infrastructure.

Given the anticipated rise in temperature extremes, especially in inland subtropical zones (Saeed et al., 2021), the increased variability could heighten these risks and intensify public health challenges in managing diarrhoeal diseases.

Climate change is also predicted to drive shifts in precipitation patterns, leading to more frequent droughts and altered seasonal rainfall (Trancoso et al., 2024). These changes, coupled with rising temperature variability, can directly reduce the availability of clean water, pushing communities to depend on potentially contaminated water sources, especially in regions with pre-existing water scarcity. During warmer, drier months, limited water availability can intensify contamination risks, because lower water levels concentrate pathogens in stagnant sources. This scenario heightens diarrhoea risk, especially in vulnerable populations with limited sanitation infrastructure.

Our study also highlights that higher maternal education reduces childhood diarrhoea, demonstrating an effect even at a regional scale across South and Southeast Asia. This finding suggests that maternal education, beyond individual households, can serve as a community-wide protective factor against diarrhoeal diseases in children. Educated mothers are often more knowledgeable about essential hygiene practices, disease prevention, and timely interventions such as oral rehydration therapy, which can save the lives of children experiencing episodes of diarrhoea. Several studies reinforce this link, showing that higher maternal education correlates with better health outcomes for children by fostering more effective responses to sanitation challenges, food safety, and hydration needs (Al Fidah et al., 2024; Fagbamigbe et al., 2021).

The World Health Organization has long recognised diarrhoea as a leading cause of morbidity and mortality, especially in children under five. The World Health Organization strategies prioritise enhancing water, sanitation, and hygiene (WASH) practices, promoting oral rehydration therapy, and strengthening health education and vaccination, such as those targeting rotavirus, to combat diarrhoeal diseases globally. Many countries have implemented these measures following World Health Organization guidelines. For instance, India’s *Clean India* campaign has improved sanitation access, and both Pakistan and Bangladesh have emphasised maternal education within their national health strategies. Bangladesh’s *Expanded Program on Immunisation* integrates maternal health literacy to empower mothers to adopt preventative measures against diarrhoea and other infectious diseases. Our study supports and expands on these initiatives by illustrating a direct association between maternal education and reduced diarrhoea incidence in children. Enhancing maternal education could therefore serve to combat diarrhoea-related mortality and morbidity, especially as climate change intensifies health vulnerabilities. Our findings also underscore that climate factors including annual temperature variability and precipitation influence diarrhoea transmission, further emphasising the need for climate-resilient health strategies.

Given this association, embedding maternal education more explicitly into public health policy could transform efforts to reduce diarrhoea incidence. Policies should prioritise accessible, scalable educational programs for women, especially in regions susceptible to water-borne diseases. This could include integrating health literacy and hygiene education within maternal and child health initiatives, expanding outreach to raise awareness on diarrhoea prevention and management, and incentivising educational opportunities for women. Incorporating maternal education into health and climate-resilience policies could substantially improve the adaptive capacity of communities to climate-related health risks. This approach not only empowers mothers to safeguard their children’s health against diarrhoeal diseases, it also contributes to fostering a healthier, more resilient generation prepared to navigate the increasing challenges posed by climate change.

Household population density was another influential variable affecting diarrhoea incidence at regional scale in South and Southeast Asia. In densely populated households, access to sanitation facilities and clean water might be limited. Overcrowding often leads to overuse of shared facilities like kitchens and toilets, increasing the risk of contamination and likelihood of poor hygiene practices that can increase the chances of transmitting pathogens causing diarrhoea. Indeed, the risk of infectious diseases is directly associated with household size (Saraswati et al., 2024) because human-to-human disease transmission increases due to close contact.

### Limitations

Our analysis relied on Demographic and Health Surveys data, which have several inherent limitations. First, Demographic and Health Surveys datasets are cross-sectional, capturing information at a single point in time rather than longitudinally. This design restricts our ability to establish causal relationships between predictors and short-term diarrhoea outcomes, because temporal changes and cause-effect sequences cannot be directly assessed. Additionally, our analysis might be conservative due to a potential positive bias introduced by the exclusion of children who have died due to diarrhoeal complications. The absence of these cases likely results in an underestimation of the true burden of diarrhoea.

Moreover, data on health outcomes and household practices are often based on caregivers’ self- reports, which can introduce recall bias, particularly for diarrhoea episodes that have occurred in the recent past. Such recall issues might affect the accuracy of data on diarrhoea prevalence and related variables. Variables such maternal education, sanitation practices, and wealth index, are susceptible to misclassification due to inconsistencies in survey administration and/or data quality issues. Further, there is potential for systematic differences between countries in terms of data collection and reporting, because the number of completed surveys can vary across countries. This reliance can also lead to potential mismatches of certain variables derived from surveys done at different times. Finally, we used WorldClim data, which offers a high spatial resolution, but represents climate conditions averaged over several decades. While suitable for examining long-term trends and spatial comparisons, these averaged data smooth out short-term temperature variation important for assessing acute health effects at fine scales.

Diarrhoea risk itself could be sensitive to short-term temperature extremes, such as unusually hot or cold days, which WorldClim data cannot capture.

## Conclusions

Our study contributes to the growing body of literature highlighting the need for climate-resilient health policies that integrate public health measures specifically targeting the reduction of diarrhoea. Our research highlights the importance of maternal education, water, sanitation, and hygiene infrastructure, and household density as important determinants of diarrhoea incidence in children under varying climate conditions. Our results therefore provide a framework for more comprehensive, targeted adaptation strategies that address both the environmental and social determinants of health. To advance public health resilience, governments in these regions should prioritise integrating climate resilience into health systems. Such integration could mitigate vulnerabilities at their root, reduce the incidence of preventable diseases, and promote healthier, more adaptable communities across South and Southeast Asia in response to a rapidly changing climate.

## Data availability

All relevant R code and data for the analyses and results available at doi:10.5281/zenodo.13864715.

## Data Availability

All relevant R code and data for the analyses and results available at doi:10.5281/zenodo.13864715.
The Demographic Health Survey (DHS) data used in this study is available upon request from the DHS Program, subject to approval by the data custodians. Detailed information on how to request access can be found on their official website: https://dhsprogram.com/data/.
The environmental data utilized in the analysis is freely accessible from WorldClim, a global climate database. It can be downloaded directly from their website: https://www.worldclim.org/data/index.html#google_vignette.

https://www.worldclim.org/data/index.html#google_vignette

## Acknowledgements

We acknowledge the sovereign Traditional Owners and custodians (First Nations) of the unceded lands where we live and work, including Kaurna in Tarndanya/Adelaide (S.H.F, C.J.A.B.), and Whadjuk Nyoongar in Boorloo/Perth (M.A.J., P.N.L.).

## Author contributions

S.H.F. and C.J.A.B did the analysis and wrote the first draft of the paper. M.A.J., P.N.L., and C.J.A.B. obtained funding. All authors reviewed, edited and provided contributions to the final manuscript.

