## Supplementary Information for "Impact of climate change on diarrhoea risk in low- and middle-income countries"

**List of tables and figures**

- **Appendix A1: Preliminary data recoding, processing, variable selection and summary statistics**
- **Table A1-1:** Preliminary data recoding, processing, variable selection and summary statistics
- **Table A1-2:** Finalised variable list with codes, recoded values, variable type specifications, and missingness
- **Table A1-3:** Descriptive statistics of diarrhoea cases among children < 5 years old at the household level.
- **Figure A1-1:** Histograms of continuous variables with missing data: comparison before and after imputation.
- **Figure A1-2:** Histograms of categorical variables with missing data: comparison before and after imputation.
- **Figure A1-3:** Correlation plots of socioeconomic variables for identifying and excluding highly correlated factors.
- **Figure A1-4:** Correlation plots of maternal traits for identifying and excluding highly correlated variables.
- **Figure A1-5:** correlation plots of child traits for identifying and excluding highly correlated variables.
- **Figure A1-6:** correlation plots of bioclimatic variable for identifying and excluding highly correlated variables.
- **Figure A1-7:** Variance plots of bioclimatic variables to identify high-variance factors.

**Appendix A1: Preliminary data recoding, processing, variable selection and summary statistics**

**Table A1-1**: Selected countries for analysis, detailing survey availability and GPS data inclusion. We used all surveys from 2010 onwards in this study.

| **Country** | **Surveys** | **GPS** |
| --- | --- | --- |
| Afghanistan | no data | no data |
| Bangladesh | **2017–2018**, **2014**, **2011**, 2007, 2004 | available |
| Cambodia | **2021–2022**, **2014**, **2010**, 2005, 2000 | available |
| India | **2019–2021**, **2015–2016** | available |
| Indonesia | 2002-2003 | available |
| Maldives | no data | no data |
| Myanmar | **2015–2016** | available |
| Nepal | **2022**, **2016**, **2011**, 2006, 2001 | available |
| Pakistan | **2017–2018**, 2006-2007 | available |
| Philippines | **2022**, **2017**, 2008, 2003 | available |
| Sri Lanka | no data | no data |
| Thailand | no data | no data |
| Timor-Leste | **2016**, **2009–2010** | available |

**Table A1-2**: Finalised variable list with codes, recoded values, variable type specifications, and missingness.

| **Variable** | **Variable Description** | **Variable code in DHS** | **Recoded values used in the final analysis** | **Variable Type** | **Missing data (%)** |
| --- | --- | --- | --- | --- | --- |
| *case ID* | case identification is used to uniquely identify each respondent | CASEID | numeric values |  |  |
| *birth sequence* | birth order index numbers the entries in the birth history from 1 to n, where nth is the first birth | BORD | numeric values |  |  |
| *country code* | alphabetic country code to identify the survey | V000 | PK7 BD7 BD6 KH8 KH6 KH5 IA7 IA6 MM7 NP8 NP7 NP6 PH8 PH7 TL7 TL5 |  |  |
| *cluster number* | cluster number | V001 | numeric values |  |  |
| *household number* | household number | V002 | numeric values |  |  |
| *respondent’s line number* | respondent’s line number | V003 | numeric values |  |  |
| **variable of interest** | | | | | |
| *diarrhoea* | child has diarrhoea in last 2 weeks | H11 | 0: no  1: yes |  | 14.54758 |
| **socioeconomics** | | | | | |
| *HH_members* | total household members | V136 | numeric values of number of members in the household | continuous | 0 |
| *household_head_sex* | sex of the head of the household | V151 | 0: male  1: female | binary | 0 |
| *wealth_index* | wealth index | V190 | 1: lowest  2: 2^nd^-lowest  3: middle  4: 2^nd^ highest  5: highest | categorical | 0 |
| *drinking_water* | main source of drinking water for members of the household | 1: improved drinking water (V113: 11, 12, 13, 14, 21, 31, 41, 51,61,62,71)  2: unimproved drinking water (V113: 32,42, 43, 44, 63, 92, 96, 97, 72)  guidelines:  dhsprogram.com/data/guide-to-dhs-statistics/household_drinking_water.htm | 0: unimproved  1: improved | binary | 2.530 |
| *sanitation_facility* | type of toilet facility in the household | derived form two variables V160 (toilet facility shared) and V116 (type of toilet facility)  1: improved sanitation facility: (V160: 0) and (V116: 11, 12, 13, 21, 22, 41)  2: unimproved sanitation facility: (V160: 1, 7) and (V116: 14, 15, 23, 31, 42, 43, 96, 11, 12, 13, 21, 22, 41)  guidelines:  dhsprogram.com/data/guide-to-dhs-statistics/type_of_sanitation_facility.htm | 0: unimproved  1: improved | binary | 0.001 |
| *displace_status* | number of years respondent has lived in the village, town, or city where interviewed | derived from V104:  value ≤ 1 ~ yes  value > 1 ~ no | 0: no  1: yes | binary | 8.96 |
| *access_healthcare* | access to healthcare facilities | derived from: V467b, V467c, V467d, V467f  0: poor access - if any of (V467b,v467c,  V467d, V467f = 1)  1: good access – all other values | 0: no  1: yes | binary | 2.73 |
| *location* | de jure type of place | V140 | 0: rural  1: urban | binary | 0.05 |
| *women_education* | respondent's education | V106: this is a standardised variable providing level of education in the following categories: no education, primary, secondary, and higher. | 0: no education  6: primary education  12: secondary education  15: higher education | continuous | 0 |
| **mother traits** | | | | | |
| *women_bmi* | body mass index for the respondent | V445 | bmi with two implied decimal points, not adjusted for pregnant women | continuous | 9.17 |
| *Women_age* | respondent's age | V012 | current age in completed years estimated from date of birth (V011) and the date of interview (V008) | continuous | 0 |
| *delivery_by_csection* | delivery by c-section | M17 | 0: no  1: yes | binary | 3.94 |
| *postnatal_checks* | baby postnatal checks | M70 | 0: no  1: yes | binary | 27.02 |
| *place_of_delivery* | place of delivery of child | M15:  place of delivery:  Public sector (20:29)  Private medical sector (30:49)  At home/other (11,12, 96)  dhsprogram.com/data/Guide-to-DHS-Statistics/Place_of_Delivery.htm | 1: Public sector  2: Private medical sector  3: At home/other |  | 5.35 |
| **child traits** | | | | |  |
| *child_gender* | sex of the child | B4 | 0: male  1: female | binary | 0 |
| *anaemia_level* | anaemia level of child measured in g/dl | HW57  anaemia levels below 7.0 g/dl are considered as severe anaemia, levels between 7.1g/dl and 9.9g/dl are considered as moderate anaemia and cases between 10.0 g/dl and 10.9 g/dl are considered as mild anaemia.  1: Not anaemic (HW57=4)  2: mild anaemia (HW57=3)  3: moderate anaemia (HW57=2)  4: severe anaemia (HW57=1)  99: missing or unknown | 0: no anaemia  1: child anaemic | binary | 20.39 |
| *HW70 (stunting)* | height for age standard deviation - stunting | HW70 | numeric values  NA: > 9000 | continuous | 31.31 |
| *HW71 (underweight)* | weight for age standard deviation - underweight | HW71 | numeric values  NA: > 9000 | continuous | 13.40 |
| *HW72 (wasting)* | weight for height standard deviation - wasting | HW72 | numeric values  NA: > 9000 | continuous | 13.00 |
| *drugs_intestinal_parasites* | drugs for intestinal parasites in last 6 months | H43 | 0: no  1: yes | binary | 17.43 |
| *birth_size* | size of the baby at birth as reported by the mother | M18: coded values were categorised as:  1 very large  2 larger than average  3 average  4 smaller than average  5 very small | 1: small  2: average  3: large | categorical | 8.20 |

**Table A1-3:** Descriptive statistics of diarrhoea cases among children < 5 years old at the household level.

| **population characteristics** | **total diarrhoea cases (*n*)** | **total children (*n*)** | **diarrhoea prevalence (%)** |
| --- | --- | --- | --- |
| **child gender** | | | |
| female | 22703 | 286262 | 8.03 |
| male | 26264 | 307053 | 8.67 |
| **age categories** | | | |
| infants (≤ 5 months) | 4860 | 55162 | 8.81 |
| babies (6–11 months) | 8304 | 58545 | 14.20 |
| toddlers (12–17 months) | 7750 | 59064 | 13.10 |
| young children (18–23 months) | 6262 | 56248 | 11.10 |
| children 24–35 months | 9317 | 115815 | 8.05 |
| preschoolers 36–47 months | 6858 | 119231 | 5.75 |
| early school age (48–60 months) | 5616 | 129250 | 4.62 |
| **countries** | | | |
| Bangladesh (BD) | 1178 | 24679 | 4.85 |
| India (IA) | 37834 | 478289 | 8.02 |
| Cambodia (KH) | 2541 | 23114 | 11.10 |
| Myanmar (MM) | 550 | 4687 | 12.00 |
| Nepal (NP) | 1537 | 15286 | 10.20 |
| Philippines (PH) | 1130 | 18749 | 6.08 |
| Pakistan (PK) | 2107 | 12094 | 17.60 |
| Timor-Leste (TL) | 2090 | 16417 | 12.9 |
| **region*** | | | |
| rural | 35433 | 426828 | 8.41 |
| urban | 10960 | 138234 | 8.04 |

*Some participants were excluded because they were not members of the household.

**Figure A1-1:** Histograms of continuous variables with missing data: comparison before and after imputation.

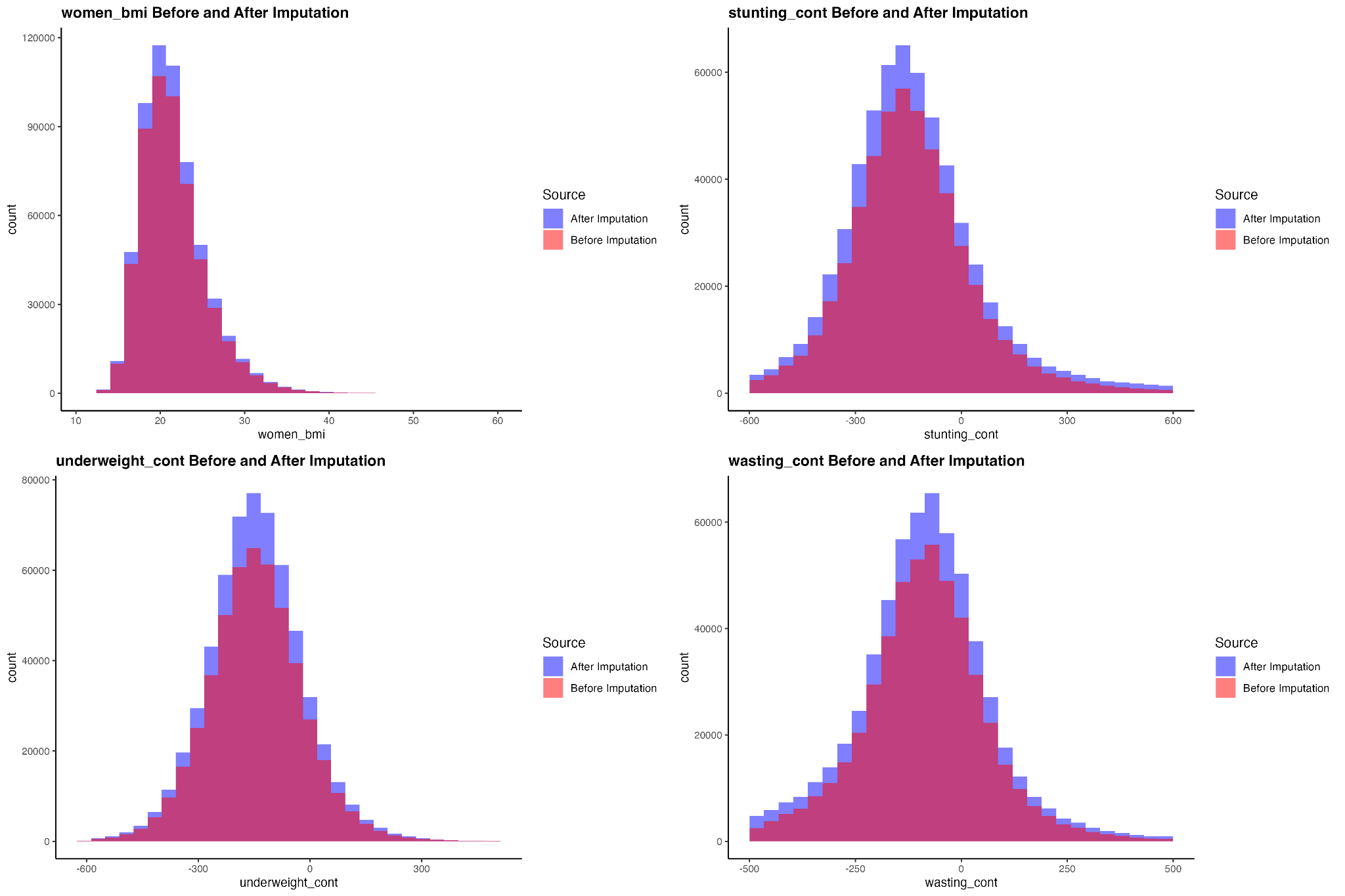

**Figure A1-2: Histograms of categorical variables with missing data: comparison before and after imputation.**

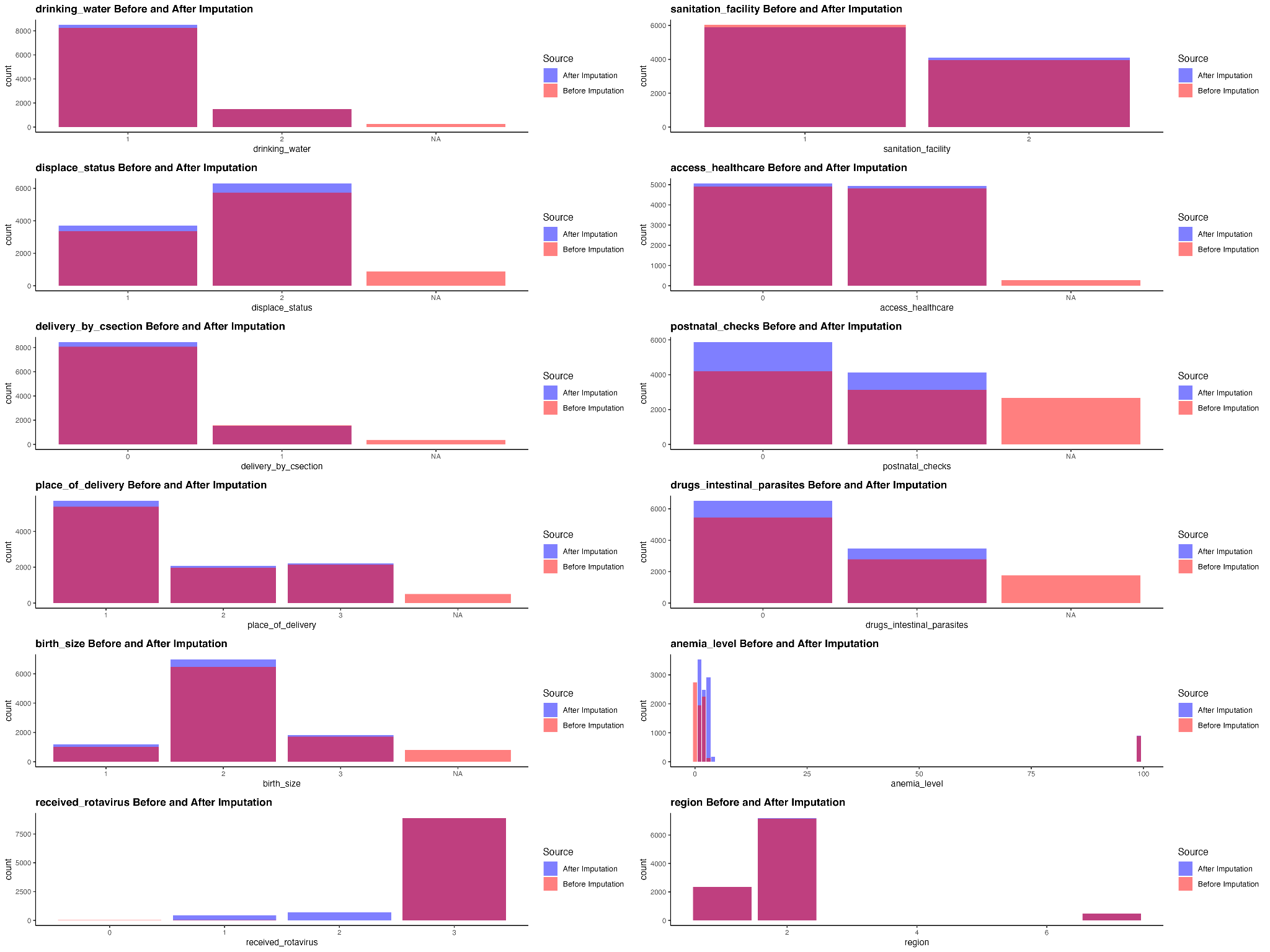

**Figure A1-3:** Correlation plots of socioeconomic variables for identifying and excluding highly correlated factors.

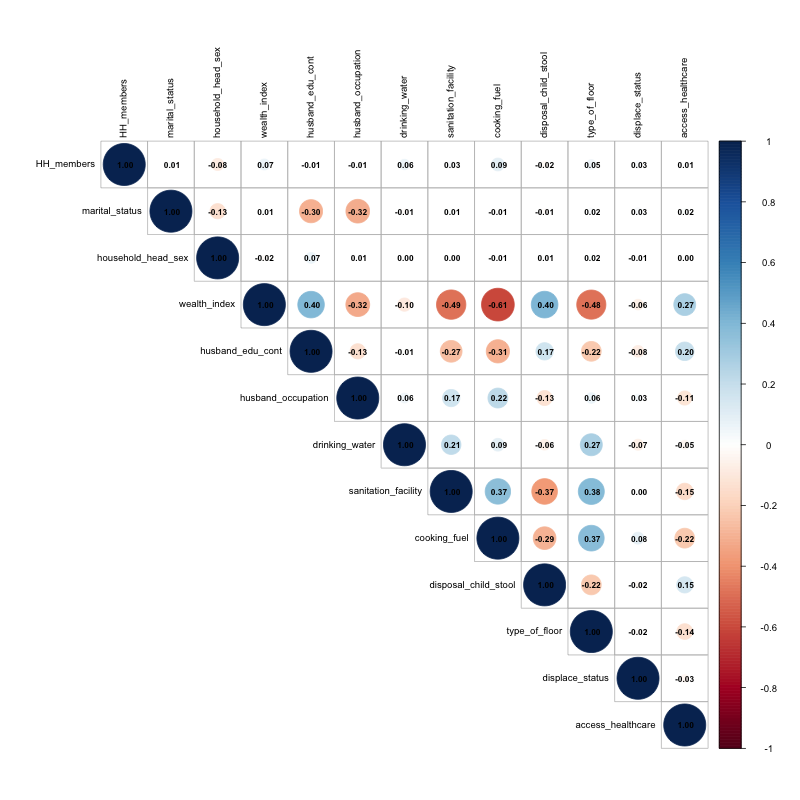

**Figure A1-4:** Correlation plots of maternal traits for identifying and excluding highly correlated variables.
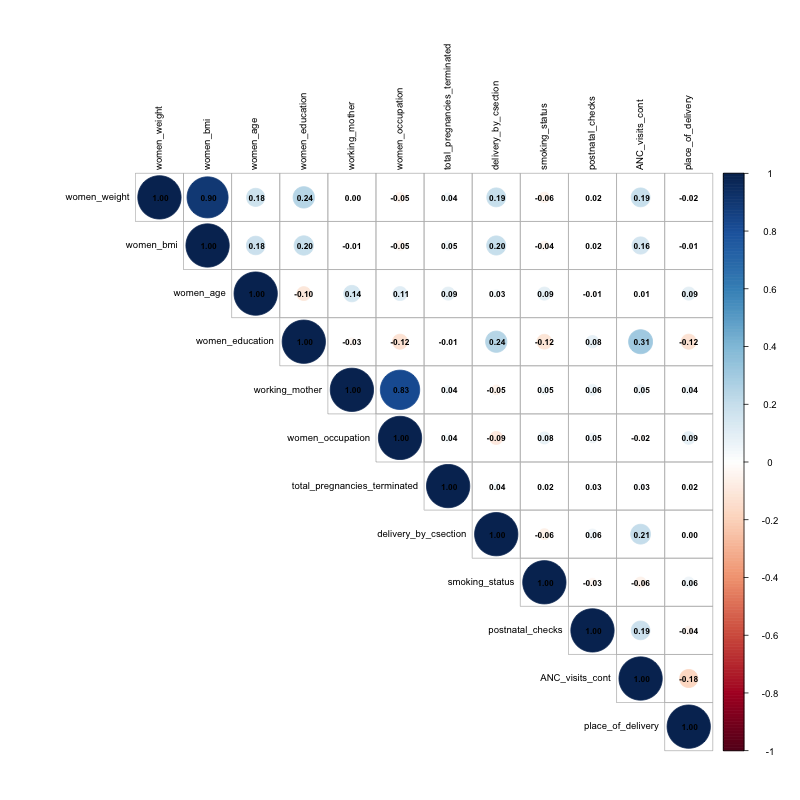

**Figure A1-5:** Correlation plots of child traits for identifying and excluding highly correlated variables.

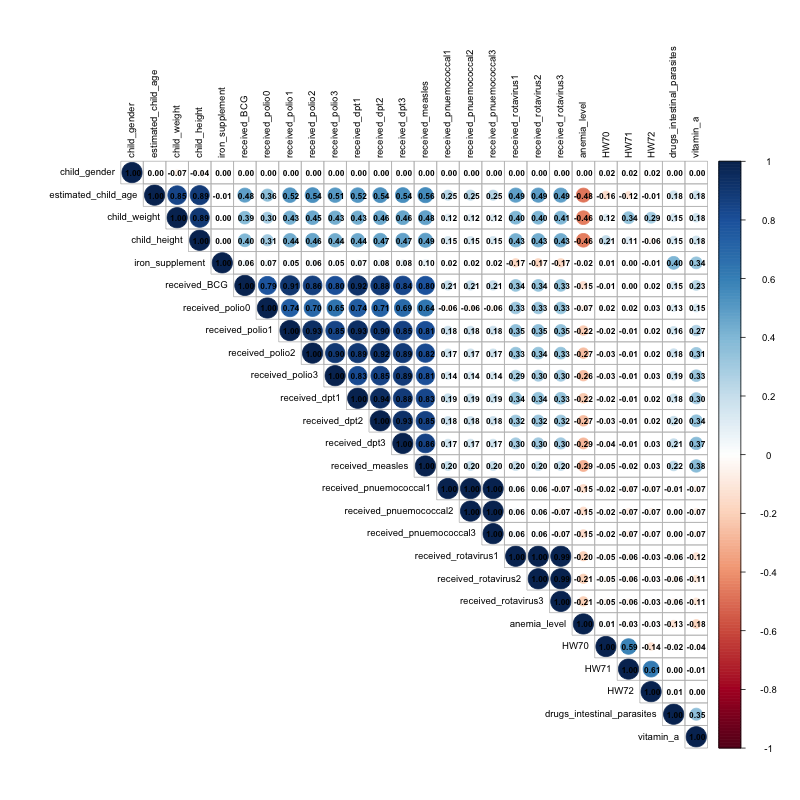

**Figure A1-6:** Correlation plots of bioclimatic variable for identifying and excluding highly correlated variables.

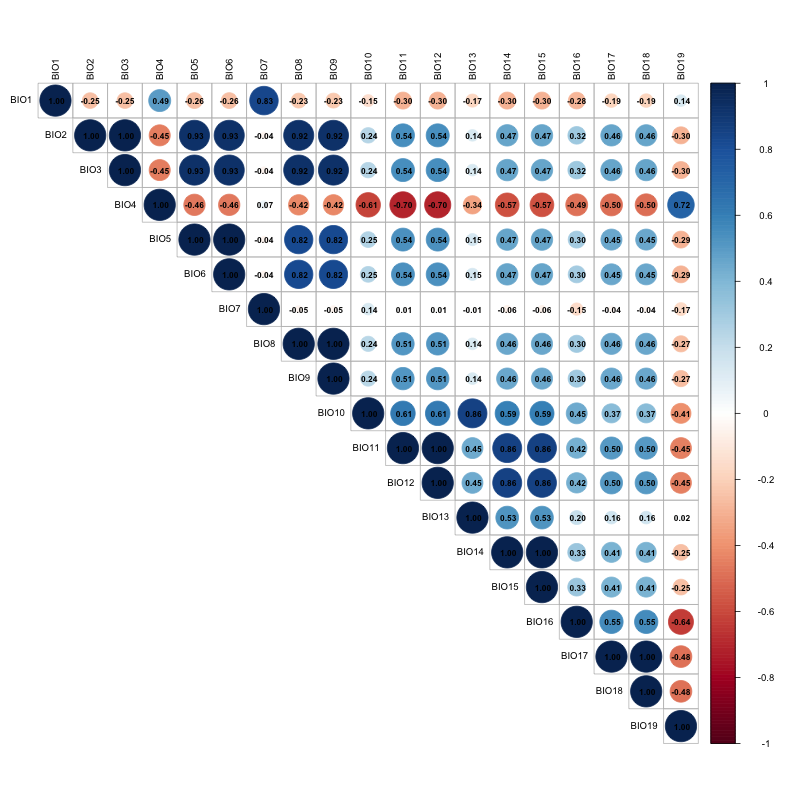

**Figure A1-7:** Variance plots of bioclimatic variables to identify high-variance factors.

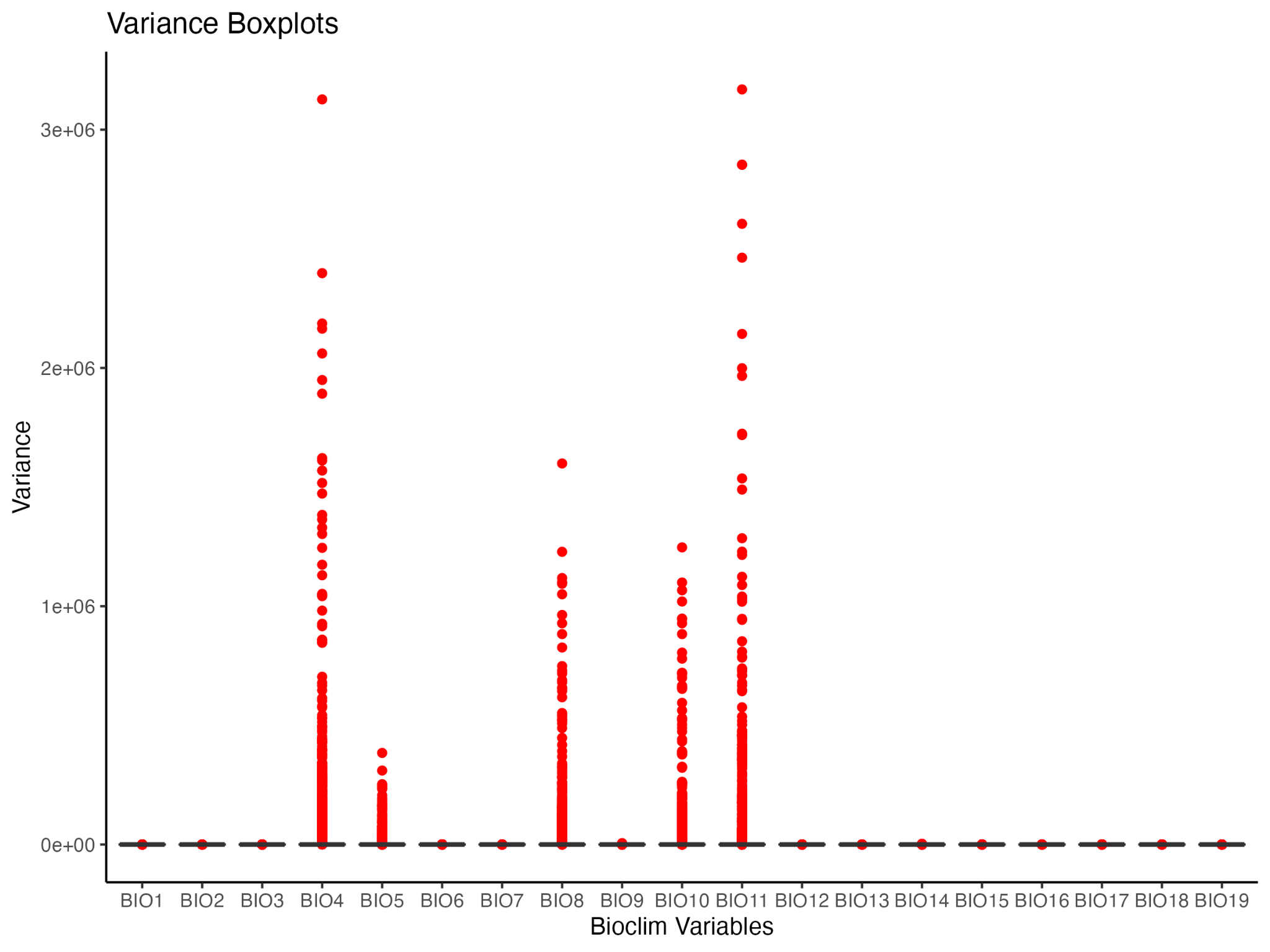

**Appendix A2: India-only analysis**

Most (56,246 of 64,843 non-missing-data clusters, or 86.7%) of the cluster data were situated in India (Fig. 1). We randomly resampled 2000 clusters (out of 56,246 total clusters, or 3.6%) per iteration (1000 iterations), demonstrating poorer overall fits (coefficients of variation: phase 1: 1.76–8.44%, phase 2: 1.09–6.53%, phase 3: 0.95–5.67%, phase 4: 4.65–12.48%, combined phase: 3.22– 11.29%) to the data relative to the full dataset, but results are approximately the same.

**Figure A2-1**: Relative influence of predictor variables on diarrhoea distribution across different model phases for India. (a) Socioeconomic factors, (b) maternal traits, (c) child traits, (d) climate variables, and (e) combined influence of the most influential variables from all phases.

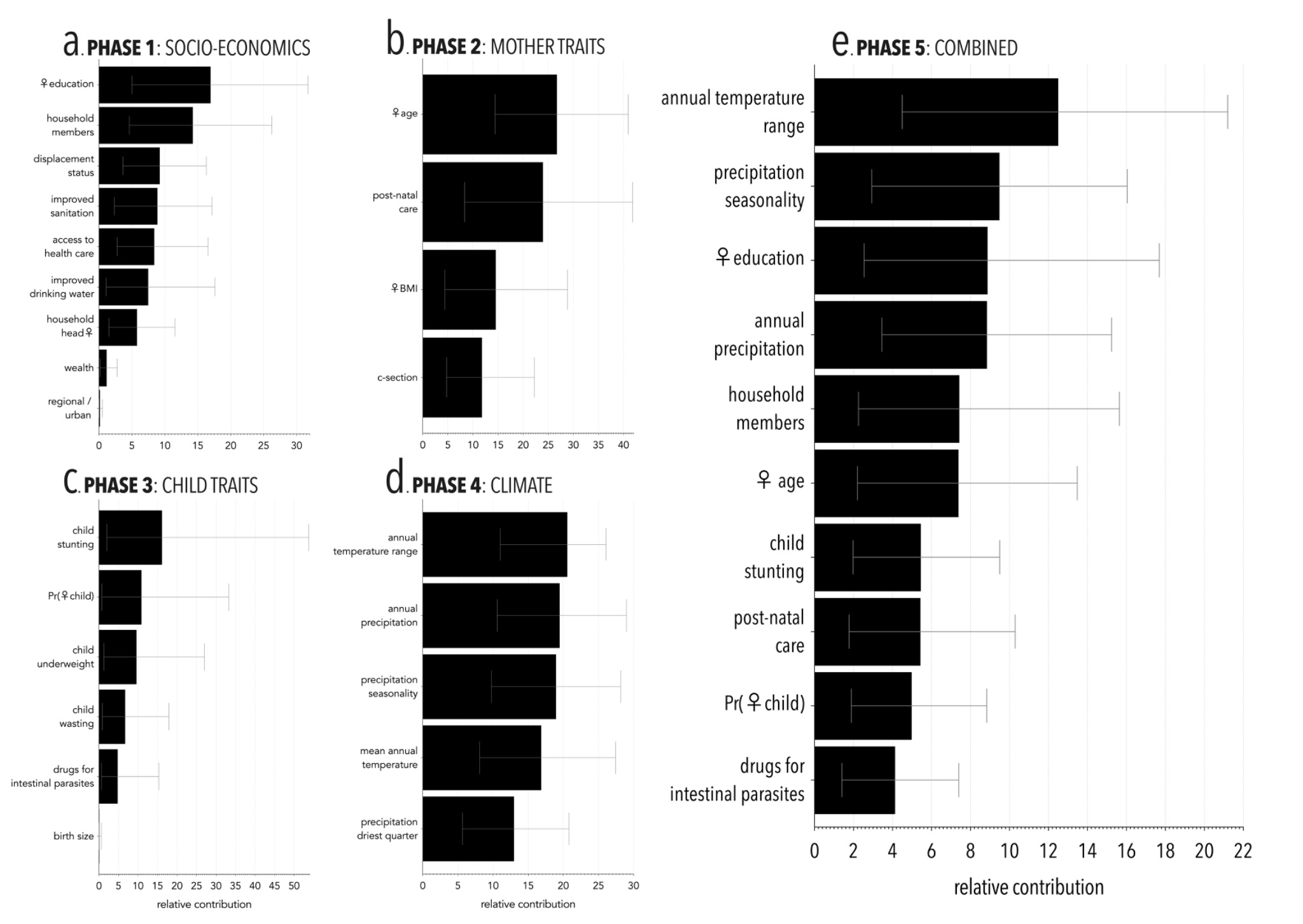

**Figure A2-2**: Predicted relationships between (a) temperature annual range, (b) precipitation seasonality, (c) mother’s education, (d) annual precipitation, (e) number of household members, (f) mother’s age on the probability of diarrhoea.

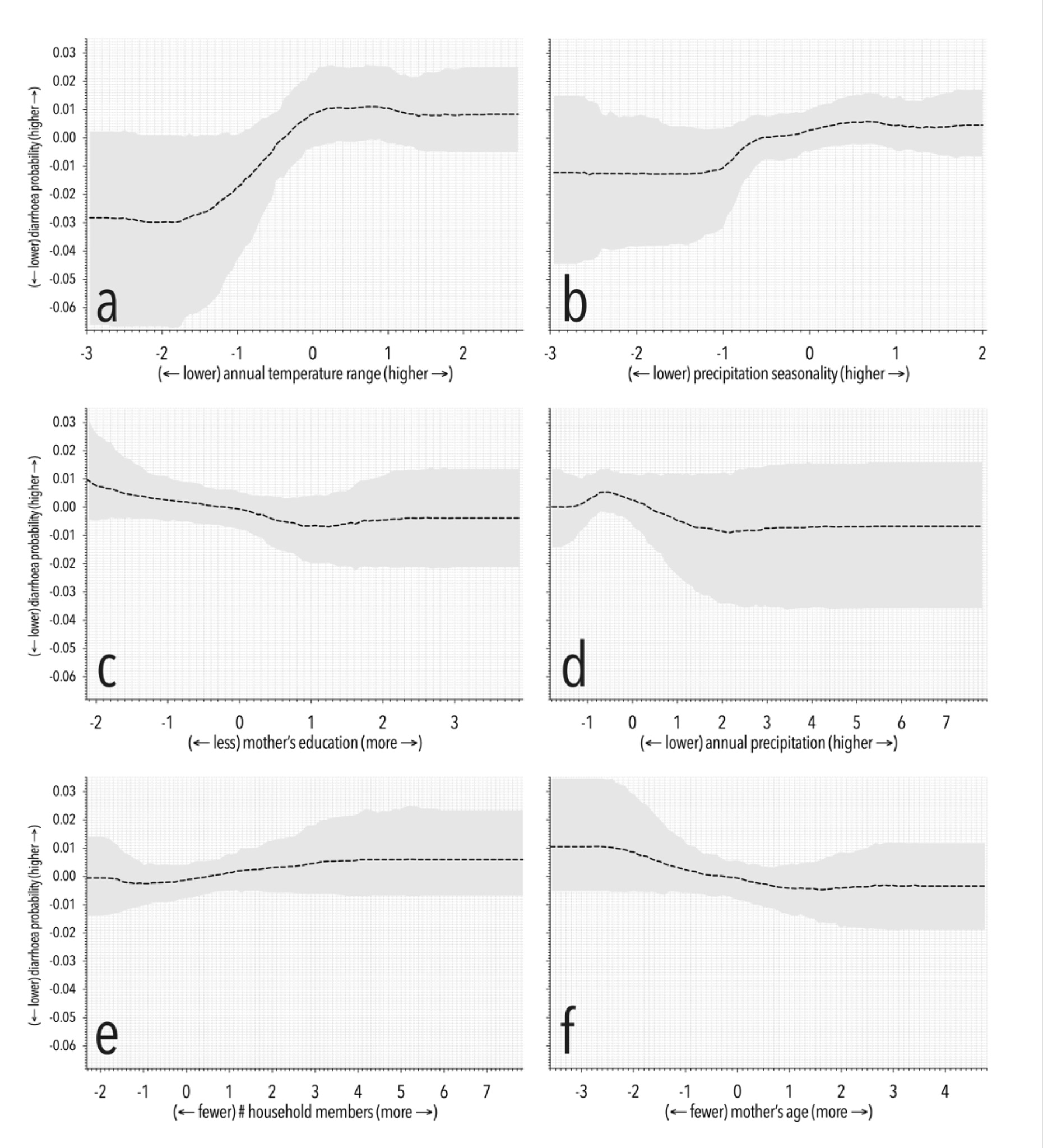
